# Epidemic analysis of COVID-19 Outbreak and Counter-Measures in France

**DOI:** 10.1101/2020.04.27.20079962

**Authors:** Eren Unlu, Hippolyte Léger, Oleksandr Motornyi, Alia Rukubayihunga, Thibaud Ishacian, Mehdi Chouiten

## Abstract

The COVID-19 pandemic has triggered world-wide attention among data scientists and epidemiologists to analyze and predict the outcomes, by using previous statistical epidemic models. We propose to use a variant of the well known SEIR model to analyze the spread of COVID-19 in France, by taking in to account the national lockdown declared in March 11, 2020. Particle Swarm Optimisation (PSO) is used to find optimal parameters for the model in the case of France. We propose to fit the model based only on the number of daily fatalities, where an *R*^2^ score based error metric is used. As the official number of confirmed cases is not reliable due to the lack of widespread testing, especially in the first phases of the outbreak, we show that basing the model optimisation on the number of fatalities can provide legitimate results.

## I. INTRODUCTION

In December 2019, novel coronavirus-sourced atypical pneumonia cases were reported in Wuhan, China. Rapidly, it evolved into an epidemic in its city of origin [1]. Despite taken counter-measures, the outbreak has gradually spread out globally. The World Health Organisation (WHO) declared it a pandemic in March 11, 2020 and called for augmented enforcing policies to all governments [2]. Countries have responded to outbreak with varying degrees of containment and other preventive actions; also with varying latency [3] [4] [5] [6]. The pandemic is ongoing by the date April 22, 2020 when this paper is written, with total confirmed cases more than 2.5 million and 180,000 deaths.

This work focuses on France, where the first confirmed case was reported on January 24, 2020. The situation then rapidly deteriorated, leading authorities to execute more draconian policies [7] [8]. A nation-wide, strict lockdown has been initiated on March 17 and was announced to continue till May 11. By April 22, there are around 159,000 confirmed cases and 20,800 deaths caused by COVID-19. French government has announced that [9] [10] lockdown will be lifted starting on May 11, with strict measures. The exact details of the lockdown lifting policy are not publicized yet, however it is known that the allowed proportion of population and the commercial and public establishments will be gradually increased week by week. The reopening date of high risk establishments such as restaurants or schools is still unknown. It would not be speculative to state that the reproduction number *R*_0_ will also increase gradually over this period, however we expect that it will be much lower compared to pre-lockdown values due to increased awareness and public health measures.

Considering this setting, we aim to analyze the current situation and its future evolution using a variation of a widely employed mathematical epidemiology model: the SEIR (Susceptible, Exposed, Infected, Recovered) model [11] [12]. We have made the assumption to use two different reproduction numbers: 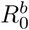 and 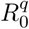. 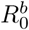 is used to describe reproduction rate between the first confirmed death until the start of the lockdown and 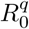 from the start to the end of the lockdown. The parameters of the model are fitted using Particle Swarm Optimization (PSO) [13] [14]. However, unlike other approaches we have decided to base our optimization on the number of fatalities only. In addition, rather than Mean Squared Error (MSE) or Mean Squared Logarithmic Error (MSLE); we minimize an *R*^2^ score based metric [15] for daily deaths. The rationale behind this approach is that due to largely unknown dynamics of the novel coronavirus such as degree of infectiousness, length of incubation period and limited testing capability, using confirmed cases with such limited information may highly disrupt the validity of the model. However, the number of deaths caused by COVID-19 is much more definitive, especially in the first days of the outbreak and thus expected to increase the accuracy. Note that, with this approach the initial state values are also defined in terms of proportion of fatalities, hence they also are parameters of the PSO optimization.

According to our initial results, we estimate the reproduction number at around 3.56 before quarantine and 0.74 after total lockdown, in agreement with various recent studies around the globe for different scenarii [16] [17]. Using the developed model we predict that if lockdown continues with strict measures, the total number of COVID-19 fatalities should topple below 50,000 (which is currently around 20,000) by late August, 2020; where the effects of the epidemic start to significantly diminish. As it is not possible to predict the reproduction number for the forthcoming lockdown lift, we propose two scenarii with a reproduction number increase of respectively 5% and 10% per week during 3 consecutive weeks. For these two scenarii, it is estimated that total number of fatalities may reach up to 70,000-80,000, and that an epidemic situation could continue till November, 2020.

## II. RELATED WORK

The SEIR model has been one of the keystone components of statistical epidemiology for a long time, and has proven its validity also for relatively recent regional or global epidemics such as MERS, SARS and Ebola [18] [19] [20], The SEIR model is classified as a *closed dynamical epidemiological model*, as it divides the total population into four distinct categories, where the proportion of individuals belonging to each category evolves over time, subjects passing from one state to other. The temporal transitions between states are defined by several differential equations [11]. Initially, the entire population is considered as susceptible except very few infected individuals. Gradually over time, according to base differential equations, susceptible individuals are exposed to disease, proportional to the reproduction number *R*_0_. Exposed subjects get infected, recover or die also based on the scalar parameters used in base differential equations. Generally, these parameters are calculated for the considered case using optimization algorithms [21], In the past, researchers have also developed noteworthy number of extended SEIR/SIR models, where additional number of states are added, considering the special circumstances of the evaluated epidemic [18] [22], We further explain the statistical formulation of an extended SEIR model in the next section.

COVID-19 pandemic has immediately gained attention among the research community and numerous different approaches using SEIR model have been proposed. First studies were published in January, 2020, for the initial epicenter Wuhan, less than a month after the novel coronavirus was identified [23] [24], One particular work aims to extend the model according to characteristics of COVID-19, by a research group from the University of Basel [25] [26]. Authors identify 3 new states: H (Hospitalized), C (Critical) and D (Dead). We base our methodology by following this guideline, as it appears to fit the dynamics of the on going pandemic.

## III. PROPOSED METHODOLOGY

As mentionned in the previous section, we developed a SEIR-HCD model as in [25]. The state transition diagram is shown in Fig. 1. Infected individuals may be hospitalized after a certain period. A proportion of the infected agents turn into critical cases, requiring intensive care; whilst the rest recovers. Among the critical cases, a certain proportion of individuals eventually dies. These proportion constants are among the parameters of the model to be optimized.

**Fig. 1.**
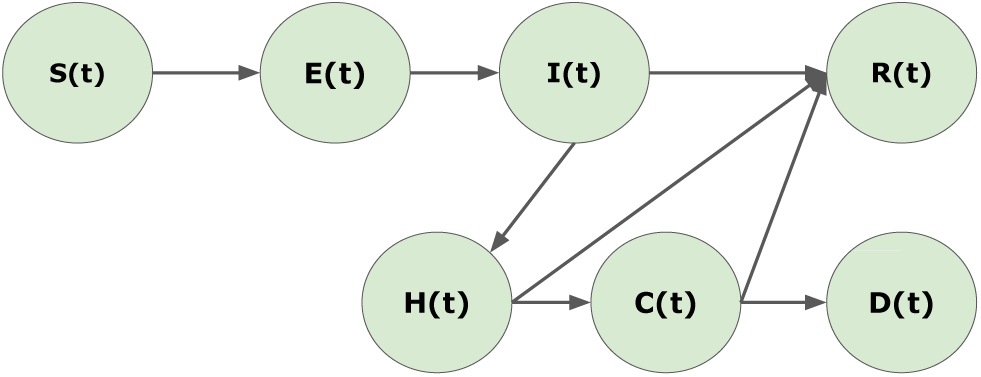
Dynamical state transition diagram of the SEIR-HCD model. Infected individuals may be hospitalized after a certain period. A certain proportion of infected agents switches to the critical case state, requiring intensive care; whilst the rest recovers. Among the critical cases, a proportion of individuals eventually die.

Initially there are only a few infected agents, *I*(*t*_0_), while rest of the population is susceptible *S*(*t*_0_) = *N* − *I*(*t*_0_); where *N* is the population of France (65 millions). Also note that, at any time the sum of all 7 states must be equal to *N*.

One of the innovative proposal we make is to base our model completely on the number of deaths. As mentioned previously, the most definitive data for COVID-19 case is the number of fatalities, due to current lack of pathological and epidemiological information about the disease and the low number of tests. Especially, in the first phase of dissemination of the virus in France, the number of tests was much lower, further decreasing the validity of using confirmed cases as a model initiator. Therefore, we propose to estimate the initial number of infected people, *I*(*t*_0_) from the initial number of fatalities *D*(*t*_0_), by simply reverse tracing the state transition diagram, using the proportion of hospitalized *H*(*t*_0_) and critical *C*(*t*_0_) cases. It is important to note that these proportion constants are parameters of the SEIR-HCD model we aim to optimize along a temporal axis. In other words, one novel outcome of our proposed algorithm is to be able to intrinsically calculate the initial number of infected citizens. It is highly reasonable to indicate that the number of infected people at the time where the first confirmed cases are announced shall be exponentially higher, due to very high proportion of asymptomatic cases [27].

The transitions between states are explained by this set of differential equations in terms of proportion of the total population [25] [26]:

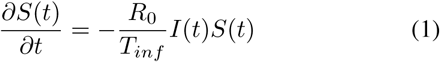

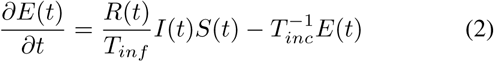

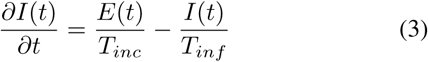

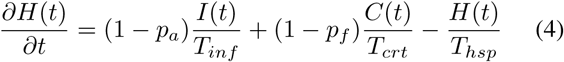

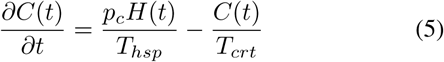

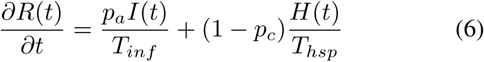

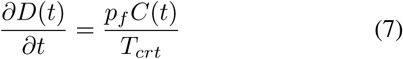

where *T_inc_* is the incubation period of the coronavirus, *T_inf_* is the infectiousness period of an infected agent, *T_hsp_* is the duration it takes for an infected agent to check in to a health facility and finally *T_crt_* is the duration it takes for an hospitalized person to turn into a critical case since the initial check-in. *p_a_, p_c_, p_f_* respectively refer to the proportion of asymptomatic infected individuals, the hospitalized agents who switched to a critical case and the critical cases resulting in death. *R*_0_ is the basic reproduction number for the coronavirus.

We have decided to use the reported data for France, starting from February 15, when the first fatality was confirmed. Without loss of generalization, we assume a binary reproduction number; 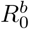 for the interval between February 15, 2020 and March 11, 2020 for the pre-quarantine period and 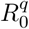 for lockdown period, which is still ongoing.

Finally, we need to optimize the model to find these 9 parameters: 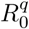, 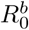, *p_a_, p_c_, p_f_, T_inc_, T_inf_, T_hsp_, T_crt_*. We employ a Particle Swarm Optimization (PSO) for this task, which is a powerful evolutionary algorithm, well suited for this setting [13].

As we propose to set our model solely on the initially reported COVID-19 related fatalities, the the initial states of each dynamic component can be denoted as:

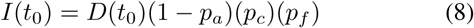

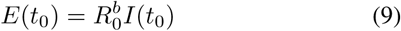

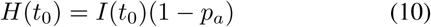

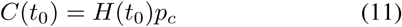

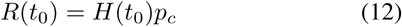

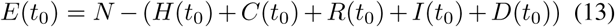

Other than suggesting a model optimization based on fatalities, we have also observed that the most adapted error metric is the *R*^2^ score of differentials of number of deaths (i. e. series of daily fatalities). This avoids overfitting, while preserving the parameter optimization in plausible ranges. The *R*^2^ score is a metric ranging from [−∞, 1], where 1 denotes the full accuracy, so we aimed to minimize the following value:

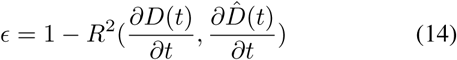

The gradual lockdown lift, starting on May 11, is modeled as a %5 to %10 increase of quarantine time reproduction number for each week for next 3 weeks (assuming reproduction number does nor grow after 3 weeks), presenting two different scenarii for France.

## IV. RESULTS

With PSO optimization based on the defined error metric, the optimal parameters for France are found in Table 1. We have estimated the reproduction number at 3.56 prior to lockdown and 0.74 after lockdown. In an independent research for France, these numbers are reported as 3.3 and 0.5, where their method is not exposed; confirming integrity of our approach from a parallel perspective [28]. We also report an approximate average reproduction number before lockdown that is consistent with the initial various studies on the issue [25] [26], where the average of the studies suggests a value around 3.6.

**Table 1.**
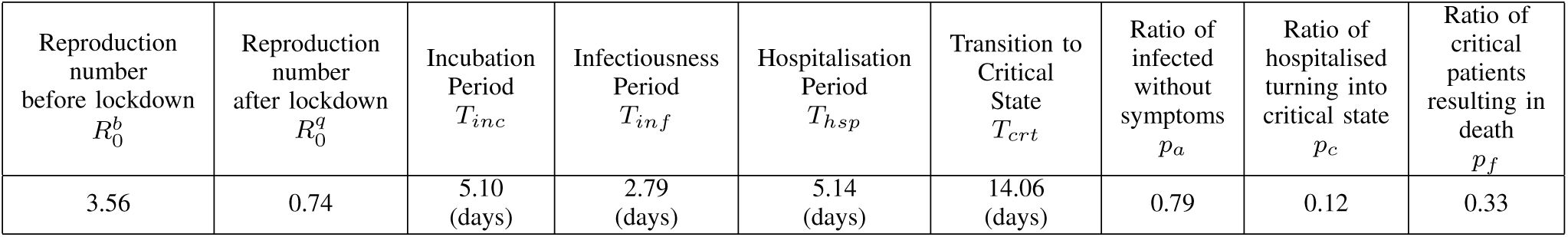
OPTIMAL VALUES OF 9 PARAMETERS OF THE SEIR MODEL FOUND BY A PSO ALGORITHM BASED ON THE *R*^2^ SCORE OF DAILY FATALITIES IN FRANCE

As shown in Table 1, all other parameters of the suggested model are within the range of other published research on COVID-19 [25]. For instance, asymptomatic ratio of 0.79, incubation period of 5.10 days, infectiousness period of 2.79 days, hospitalisation period of 5.14 days, transition to critical state period of 14.06, ratio of 0.12 for infected becoming critical and fatality ratio of 0.33 for critical patients are all close to the medians of the other reported work [25]. These results are particularly interesting, since our SEIR model optimization approach with suggested error metric for number of fatalities is able to converge to the optimal point, and directly coronavirus related metrics such as incubation period, hospitalisation period etc. (assuming demographics etc. of a country does not influence considerably) are close to the average of the previous works. This asserts the validity of binary reproduction number approach for lockdown and its calculated values. Note that, in complex dynamics of a SEIR model, slight variance of parameters may have a drastic impact on the outputs, such as the estimated number of deaths.

We have calculated the mortality ratio of infected people (1 − *p_a_*)*p_c_p_f_* as %0.83, where [28] proposes %0.5 for France. By the date April 22, our model estimates the proportion of already infected population as %6.6 (Fig. 2–3), while [28] also reports as %6. It is quite surprising to observe independent works show similar results, while our SEIR model optimization only takes fatalities into account for optimization.

**Fig. 2.**
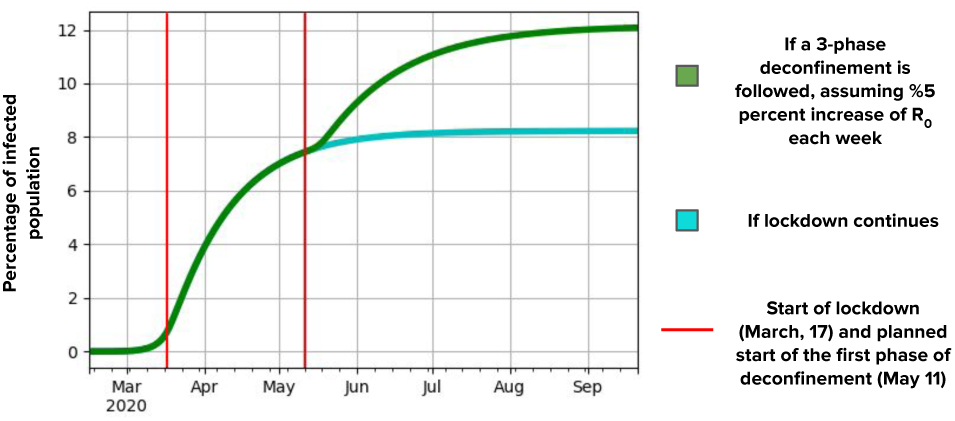
Predicted number of infected proportion of France from February 15, 2020 to September 21, 2020; considering a scenario, where quarantine time reproduction number grows by 5% each week, during 3 weeks following lockdown lift.

**Fig. 3.**
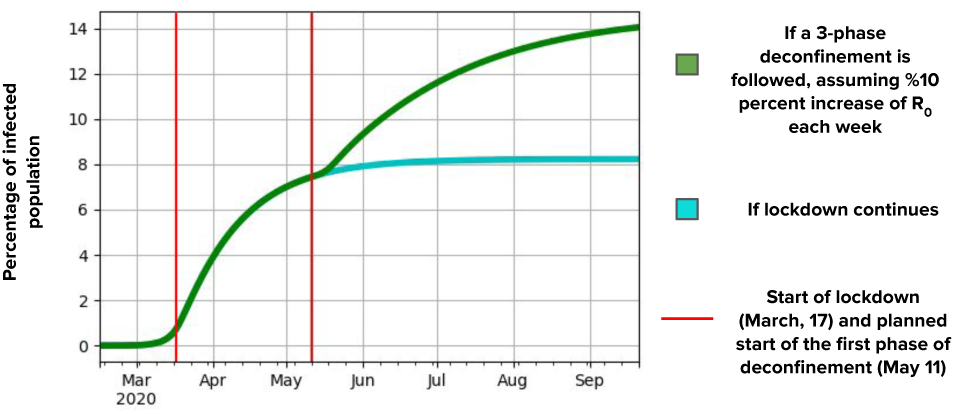
Predicted number of infected proportion of France from February 15, 2020 to September 21, 2020; considering a scenario, where quarantine time reproduction number grows by 10% each week, during 3 weeks following lockdown lift.

Based on our model, we estimate that if lockdown is maintained, the number of fatalities for France might never pass the limit of 50,000. If reproduction number grows by 5% each week during 3 weeks following the lockdown lift; this upper limit might reach 70,000 (Fig. 4). We observe that, if strict measures are respected till September, 2020 the epidemic almost terminates around this date. In case this growth rate becomes 10% per week, the upper death toll limit might reach 80,000 (Fig. 5); where epidemic situation lasts till November, 2020. Considering the fact that, French government has already prepared a versatile and extensive plan for lifting, by limiting social gatherings, augmented surveillence and nationwide distribution face masks, these scenarii with %5-%10 per week increase of quarantine time reproduction number seem legitimate.

**Fig. 4.**
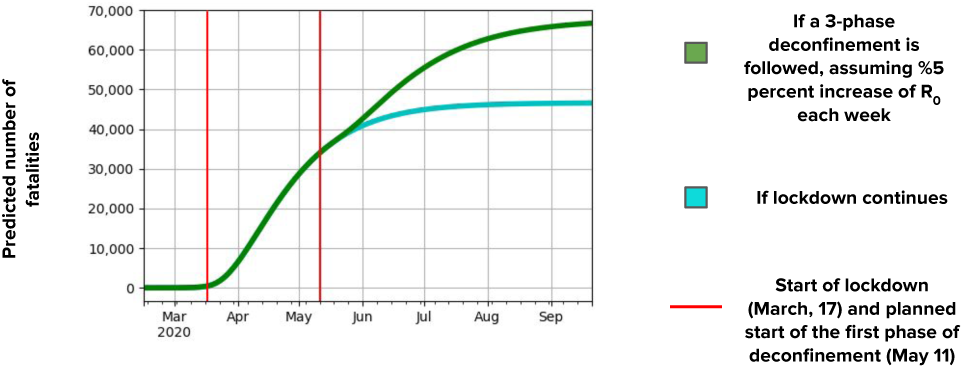
Predicted number of fatalities for France from February 15, 2020 to September 21, 2020; considering a scenario, where quarantine time reproduction number grows by 5% each week, during 3 weeks following lockdown lift.

**Fig. 5.**
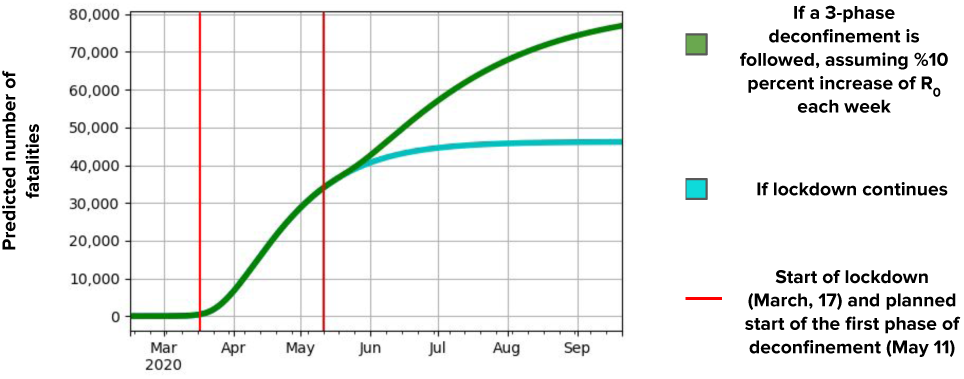
Predicted number of fatalities for France from February 15, 2020 to September 21, 2020; considering a scenario, where quarantine time reproduction number grows by 10% each week, during 3 weeks following lockdown lift.

## V. CONCLUSION

In this paper, we have proposed a SEIR model for COVID-19 epidemic in France, similar to [25]. Unlike other similar attempts, we have used only confirmed number of deaths as an optimization metric. Rationale behind this proposal is that deficiency in testing coverage, especially in the early phases of the outbreak, greatly underestimates the number of confirmed cases. However, the number of confirmed fatalities is a much more solid evidence in this setting. An error metric based on the daily death toll is presented and model parameters are optimized using PSO algorithm. Also note that, the initial state of the model (initial values of each state) is also defined based on the proportion of fatalities, hence all initial state values are also parameters for the PSO optimization. We believe that, for epidemics at this scale starting a SEIR models with a hypothetical single infected individual may greatly lower the accuracy; especially considering their potential drastic impact.

## Data Availability

All data related to coronavirus daily fatalities are taken from open sources.

https://coronavirus.jhu.edu/

